# Risk Factors of Developing Contralateral Breast Cancer After Treatment of a First Primary Breast Cancer

**DOI:** 10.1101/2022.06.30.22277087

**Authors:** Maryam Avatefi, Fatemeh Hadavand-Siri, Seyed-Saeed Hashemi-Nazari, Mohammad Esmaeil Akbari

## Abstract

**Introduction:** Breast cancer (BC) is the most common cancer among women worldwide. This study aimed to determine the disease-free interval (DFI) and the effect of risk factors and characteristics of first primary breast cancer survivors on the progression of contralateral breast cancer (CBC).

**Method:** In this retrospective cohort study, we identified 5003 women (containing 145 CBC and 4858 BC survivors) diagnosed with BC between 2000 and 2020 in the cancer research center. CBC was diagnosed at least 6 months after detection of primary BC. Kaplan–Meier method was used to indicate DFI and survival curves. To determine the risk factors of CBC, the Cox proportional hazard regression model was employed.

**Results:** The median time interval among primary BC and CBC was 7.10 years (average: 7.57, range: 0.49-19.95). The 5-year DFI was 97%. The median age of CBC and primary BC patients was 47 (range 17-78) and 48 (range 17-90), respectively. ER+, PR+, and HER2+ were reported in 72.13%, 66.67%, and 30% of CBC patients. Also, 69.57% of patients had IDC pathology type and 81.90% and 83.64% of the patients were treated with adjuvant chemotherapy and external radiotherapy. More than half of the participants had no family history of BC (69.57%). The hazard ratio indicates that women 60-70 years old, a higher number of involved lymph nodes, and recurrence had significant relationships with CBC.

**Conclusion:** This is the first study to investigate the risk factors of CBC and disease-free interval among BC survivors. Women with higher lymph node metastasis have a lower chance to develop CBC and patients with recurrence are at higher risk for CBC.

## Introduction

Worldwide, one of the increasing health problems is raising the incidence of malignancies(1). The criteria for detection of a second primary neoplasm have been studied and discussed for over a century(2). Globally, breast cancer is the most common cancer among females has 2.3 million new cases in 2020, and the female breast cancer death rate was 15 per 100,000 in transitioning countries as reported by GLOBOCAN(3).

Breast cancers compared to other cancers, have a notable association with second primary cancers and can happen as second primary cancers themselves(4). The improvement in diagnosis and treatment of breast cancer and increasing life expectancy have led to a higher risk of bilateral breast cancer (synchronous (SBC) or metachronous breast cancer (MBC))(5, 6). Metachronous malignancy is diagnosed at least 6 months after the first tumor(7). In the last 3 decades, the incidence of metachronous breast cancer in different studies has been reported in the range of 1.4%-12% with a follow-up time of 6-15 years(8, 9).

There is no strong evidence of whether contralateral development of cancer leads to poor survival in comparison with unilateral disease. This represents a relative risk two to six times higher than the Progression of the first breast cancer in the general population(10). Patients with second primary cancer have lower 5-year survival rates than those who had primary malignancies(11).

The interval between treatment of primary neoplasm after the identification of second primary cancer is called the disease-free interval (DFI). The disease-free intervals’ effect on the second primary tumor prognosis is not well defined. Based on the prior reports DFI was categorized into two groups: short and long DFI. The first five years after the primary tumors’ detection and the following years as short and long DFI, respectively. The long DFI is associated with a better prognosis and favorable outcomes(12, 13).

Second primary cancers have occurred because of environmental risk factors, genetic factors, lifestyle, or treatment-related factors (14). The most important risk factors of second primary breast cancer include reproductive factors, lobular histology, mutation of BRCA1 / BRCA2, and family history of breast cancer(15). However, breast cancer is very heterogeneous, and different risk factors can play a key role in the occurrence of second primary breast cancer, which can vary depending on the community studied in different studies.

Although many studies have been done on the risk factors and epidemiology of breast cancer, less research has been published on the related factors of second primary breast cancer. Also, a review of studies conducted in Iran shows that less information has been published about the risk factors of bilateral breast cancer after treatment of unilateral breast cancer. Therefore, considering the importance of this issue, this study aims to address the risk factors of developing contralateral breast cancer (CBC) after treatment of first primary breast cancer. We also assessed the disease-free interval of patients with first primary breast cancer. It needs to clarify the incidence, clinicopathological characteristics, and disease-free interval of contralateral breast cancer.

## Material and Methods

### Study design

In this retrospective cohort study, data of patients were obtained from the cancer research center of Shahid Beheshti University of Medical Sciences between 2000 to 2020. The cancer research center registers breast cancer patients who are followed up at least annually with any required investigations and routine clinical examinations. After obtaining permission from the ethical committee of our institute, the cancer research center of Shahid Beheshti University of Medical Sciences (No: IR.SBMU.CRC.REC.1400.048), to evaluate the risk factors of developing CBC, we studied age at first primary breast cancer, tumor size, estrogen, and progesterone receptor of the primary tumor, pathological types, lymph vascular invasion, chemotherapy, HER2 (human epidermal growth factor receptor 2) levels and history of radiotherapy, cancer recurrence, stage, grade, family history of cancer, number of lymph nodes involved in the axilla. We also, inside investigating CBC risk factors evaluate the disease-free interval which is defined as the time between diagnosis of 1st breast cancer and second primary malignancy in the other breast.

### study population

After reviewing the 5515 women with diagnosed primary breast cancer between follow-up time of study, those patients without synchronous or metachronous tumors and non-metastatic primary breast cancer at the time of admission, were selected. Bilateral synchronous breast cancers and those patients who follow less than 180 days were also a measure of exclusion from the study (N=375). Only metachronous breast cancers (metachronous tumors were defined as tumors that appear at least 6 months after the first tumor)(7), were considered (N=145). Also, we excluded patients with incomplete profiles (137 observations deleted). In total the remaining 5003 subjects were analyzed.

### Data analysis

Descriptive analysis of the data on variables was performed for total sample (n=5003) enrolled in the study, reporting the mean and standard deviation and median for continuous and frequencies for categorical variables.

The Chi-square test and t-test were used for categorical and continues variables respectively to compute different variables’ frequency among groups (contralateral and non-contralateral breast cancer patients). The Kaplan-Meier was employed to compare DFI (disease-free interval) and time to recurrence among the groups (contralateral and non-contralateral breast cancer patients). Also, the Cox proportional hazard regression model was adopted to estimate the crude and adjusted hazard ratio (HR) for the risk of contralateral breast cancer (CBC).

The Cox proportional hazards assumption was tested both numerically.

All the variables with a P-value less than 0.2 or variables that most of the levels had a P-value less than 0.2 in univariate analysis were included in the multivariable analysis to identification of related factors of CBC. A significance level of 0.05 was considered. Stata (version 14.0; Stata Corp, Texas, USA) software was used to perform the analyzes.

## Results

### Study characteristics

Regarding study design, 5003 approved unilateral breast cancer patients were studied.

Of the selected patients, according to the CBC approval and definition, 145 patients (2.9%) with a median age of 47 (range 17-78) progressed to CBC within the following years after treatment. All the other 4858 patients with a median age of 48 (range 17-90) were considered as unilateral breast cancer group. The median time interval among primary BC and CBC was 7.10 years (average: 7.57, range: 0.49-19.95). Of these, 54.51% of patients were younger than 50 years old. More than half of (57.65%) patients’ tumor size was 2-4 cm. As compared to patients with primary breast cancer, in counterparts with CBC 61.54% of tumors were 2-4 cm in size. We found 72.13% and 66.67% of subjects have ER^+^ and PR^+^, respectively but 70% of patients have negative HER2. The most common types of breast cancer histology were IDC (69.57%). For 74.63% of patient’s adjuvant chemotherapy was used. Nearly all (83.53%) of the subjects were treated with external radiotherapy and 61.22% of patients have negative lymph vascular invasion. 69.57% of patients didn’t have a family history of breast cancer (in first or second degrees). Of the patients, 51.32% haven’t any involved lymph nodes. **Table 1** contains the patient’s characteristics.

**Table 1.** Factors associated with the CBC after primary breast cancer treatment

| **Variables** | **Total (N=5003)**<br>**N (%)** | **UBC (N=4858)**<br>**N (%)** | **CBC (N=145)**<br>**N (%)** | **p-value** |
| --- | --- | --- | --- | --- |
| **Mean age at diagnosis (SD)** | 49.21±11.77 | 49.28±11.81 | 47.23±9.96 | 0.042 |
| **Median (range)** | 48 (17-90) | 48 (17-90) | 47 (17-78) |  |
| **Age** |  |  |  |  |
| <50 | 2482 (54.51) | 2404 (54.46) | 78 (56.12) | 0.03 |
| 50~ | 1190 (26.14) | 1144 (25.92) | 46 (33.09) |  |
| 60~ | 624 (13.70) | 612 (13.86) | 12 (8.63) |  |
| >70 | 257 (5.64) | 254 (5.75) | 3 (2.16) |  |
| **Tumor size (cm)** |  |  |  |  |
| < 2 | 1001 (27.00) | 978 (27.14) | 23 (22.12) | 0.52 |
| 2~ | 2137 (57.65) | 2073 (57.54) | **64 (61.54)** |  |
| >5 | 569 (15.35) | 552 (15.32) | 17 (16.35) |  |
| **ER status** |  |  |  |  |
| Positive | 3023 (74.98) | 2935 (75.06) | **88 (72.13)** | 0.46 |
| Negative | 1009 (25.02) | 975 (24.94) | 34 (27.87) |  |
| **PR status** |  |  |  |  |
| Positive | 2771 (68.71) | 2691 (68.77) | **80 (66.67)** | 0.62 |
| Negative | 1262 (31.29) | 1222 (31.23) | 40 (33.33) |  |
| **Her2 status** |  |  |  |  |
| Positive | 893 (23.09) | 860 (22.89) | 33 (30.00) | 0.08 |
| Negative | 2974 (76.91) | 2897 (77.11) | **77 (70.00)** |  |
| **Breast histology** |  |  |  |  |
| IDC | 3426 (70.73) | 3330 (70.76) | **96 (69.57)** | 0.17 |
| DCIS | 241 (4.98) | 235 (4.99) | **6 (4.35)** |  |
| ILC | 286 (5.90) | 272 (5.78) | **14 (10.14)** |  |
| Others | 891 (18.39) | 869 (18.47) | 22 (15.94) |  |
| **Chemotherapy** |  |  |  |  |
| No | 355 (9.20) | 342 (9.13) | 13 (11.21) | 0.02 |
| Adjuvant | 2883 (74.63) | 2788 (74.41) | **95 (81.90)** |  |
| Neoadjuvant | 625 (16.17) | 617 (16.47) | 8 (6.90) |  |
| **Radiotherapy (RT)** |  |  |  |  |
| No | 105 (2.77) | 100 (2.71) | 5 (4.55) | 0.49 |
| External RT | 3170 (83.53) | 3078 (83.53) | **92 (83.64)** |  |
| IORT Boost | 335 (8.83) | 325 (8.82) | 10 (9.09) |  |
| IORT Radical | 185 (4.87) | 182 (4.94) | 3 (2.73) |  |
| **lymph vascular invasion (LVI)** |  |  |  |  |
| Positive | 1417 (38.47) | 1379 (38.47) | 38 (38.78) | 0.95 |
| Negative | 2266 (61.53) | 2206 (61.53) | **60 (61.22)** |  |
| **Family history** |  |  |  |  |
| No | 2625 (72.62) | 2545 (72.71) | **80 (69.57)** | 0.11 |
| Second degree | 529 (14.63) | 505 (14.43) | 24 (20.87) |  |
| First degree | 461 (12.75) | 450 (12.86) | 11 (9.57) |  |
| **Lymph node** |  |  |  |  |
| 0 | 2135 (51.32) | 2070 (51.22) | 65 (54.62) | 0.28 |
| 1~ | 1077 (25.89) | 1055 (26.11) | **22 (18.49)** |  |
| 4~ | 631 (15.17) | 610 (15.10) | 21 (17.65) |  |
| >10 | 317 (7.62) | 306 (7.57) | 11 (9.24) |  |
| **Stage** |  |  |  |  |
| I | 886 (21.10) | 860 (21.09) | 26 (21.31) | 0.84 |
| II | 1837 (43.75) | 1781 (43.68) | **56 (45.90)** |  |
| III, IV | 1476 (35.15) | 1463 (35.22) | 40 (32.79) |  |
| **Grade** |  |  |  |  |
| I | 426 (10.89) | 413 (10.85) | 13 (12.26) | 0.88 |
| II | 2132 (54.49) | 2076 (54.53) | **56 (52.83)** |  |
| III | 1355 (34.62) | 1318 (34.62) | 37 (34.91) |  |
| **Recurrence** |  |  |  |  |
| No | 4376 (87.47) | 4271 (87.92) | **105 (72.41)** | <0.001 |
| Yes | 627 (12.53) | 587 (12.08) | 40 (27.59) |  |

### The DFI Between PBC and CBC / Recurrence

The 1-, 5-, 10-, and 20-year DFI was 99, 97, 96, and 88% in breast cancer acquired patients, respectively (**Fig. 1**). The 5-, 10-, and 15-year DFI between PBC and recurrence in patients who have CBC were significantly lower than in non-recurrence patients and the overall DFI of recurrence was 63% and 81% for CBC and non-CBC groups, respectively (**Fig. 2**).

**Figure 1.**
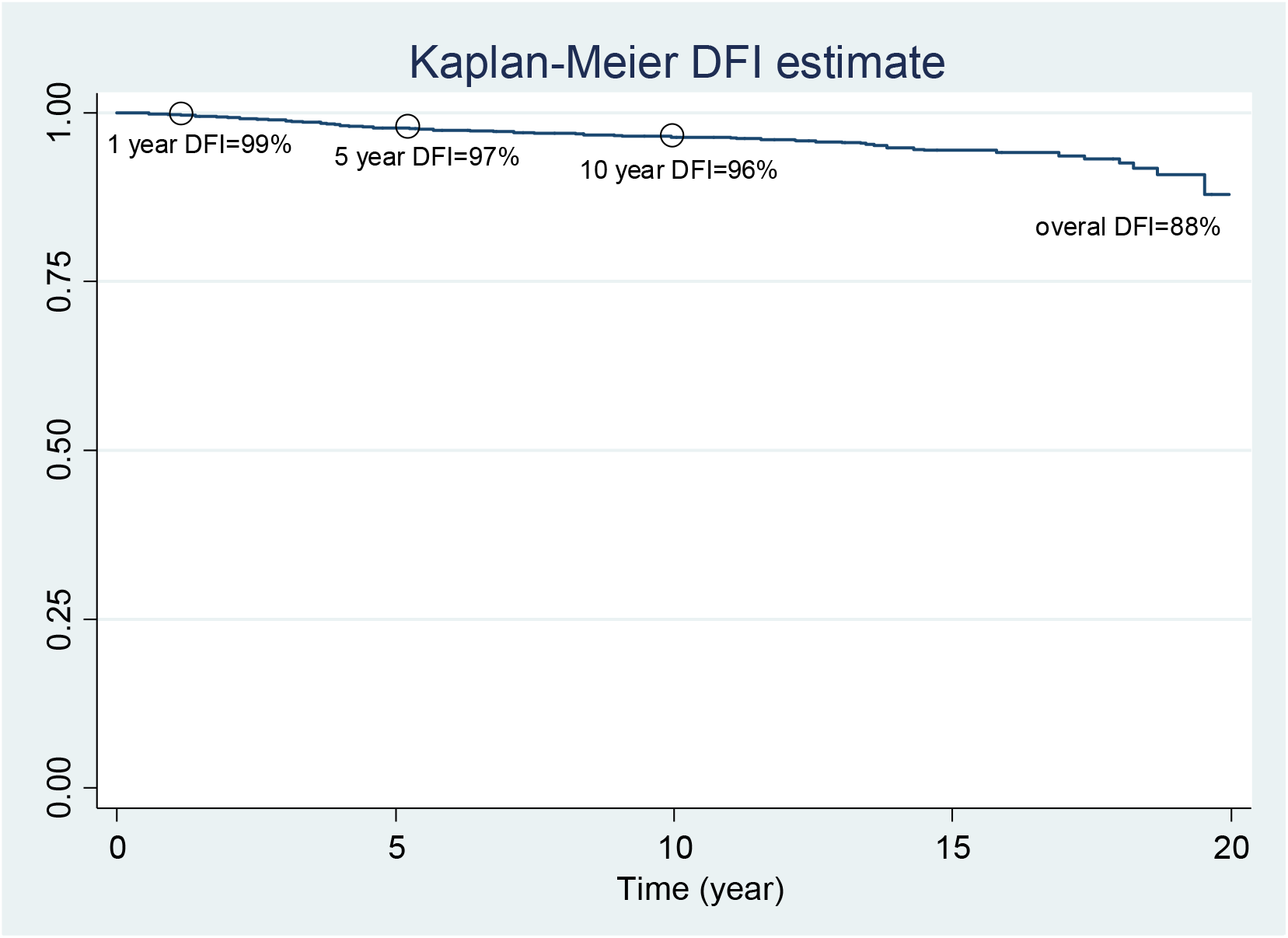
Kaplan-Meier curve of 1-, 5-, 10-, and 20-year disease-free interval for CBC after primary BC treatment

**Figure 2.**
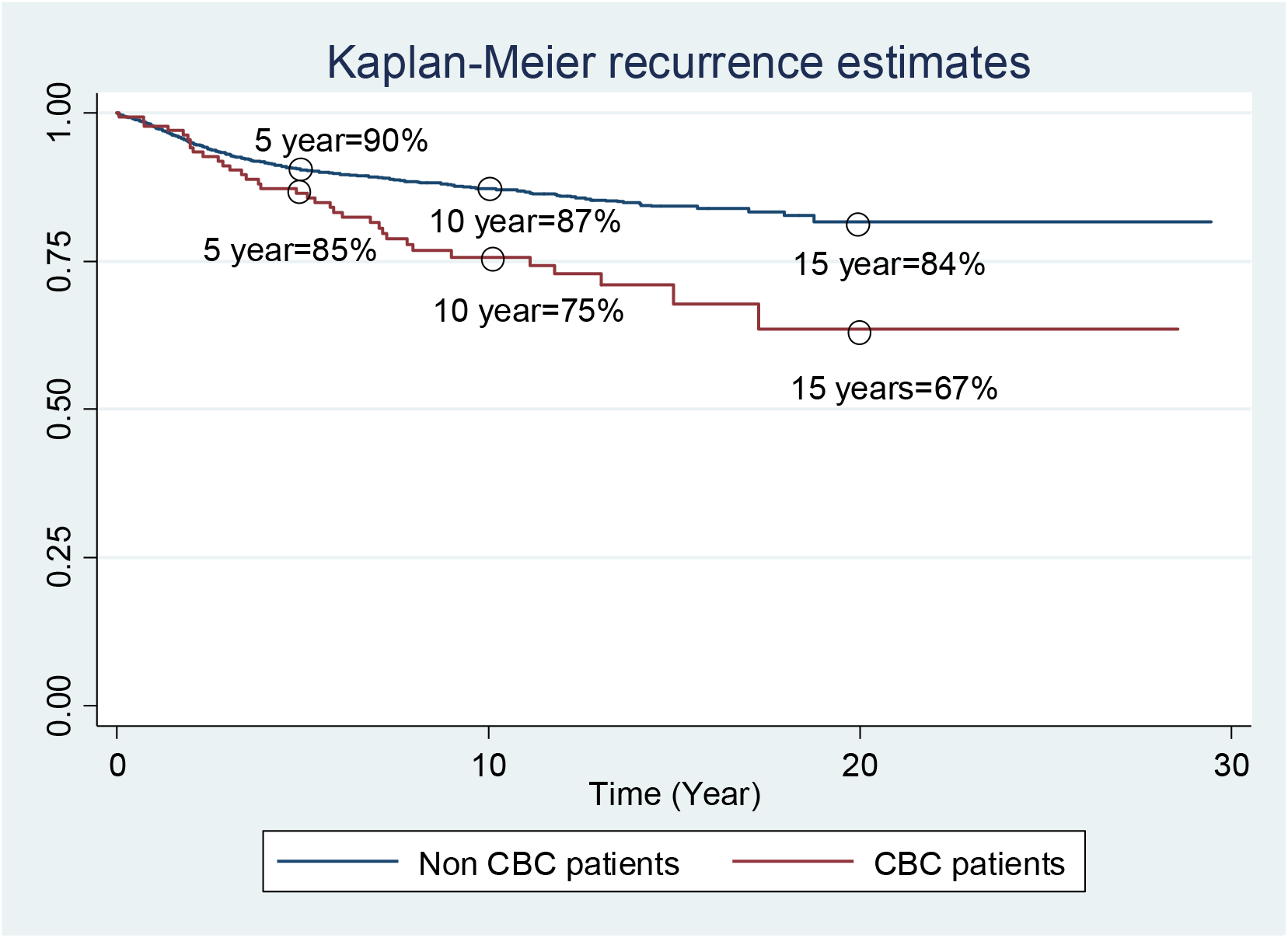
Kaplan-Meier curve of 5-, 10-, and 15-year disease-free interval of recurrence after primary BC treatment

**Table 2** contains results of univariable and multivariable Cox proportional hazards regression models. Most variables were protective. The IDC type of pathology was significantly associated with the appearance of CBC and it was a protective factor (HR = 0.61; 95% CI: 0.38–0.97). Patients who were treated with neoadjuvant chemotherapy had 60% less likely to develop second primary breast cancer (HR = 0.40; 95% CI: 0.16–0.97). Family history of breast cancer in the second degree increased the chance of CBC by 59%. Recurrence made a difference in the progression of CBC. The recurrence of primary breast cancer increased the risk of CBC (HR = 2.17; 95% CI: 1.50–3.12).

**Table 2.** Multivariable Cox regression analysis for CBC risk factors

| Variables | Hazard Ratios (95% confidence intervals) |  |
| --- | --- | --- |
|  | Univariable model<br>HR (95% CI) | Multivariable model<br>HR (95% CI) |
| **Age** |  |  |
| <50 | Ref. | Ref. |
| 50~ | 1.18 (0.82 to 1.70) | 0.99 (0.61 to 1.59) |
| 60~ | 0.63 (0.34 to 1.15) | 0.32* (0.11 to 0.89) |
| >70 | 0.35 (0.11 to 1.12) | 0.54 (0.16 to 1.78) |
| **Tumor size (cm)** |  |  |
| < 2 | Ref. | - |
| 2~ | 1.18 (0.73 to 1.91) | - |
| >5 | 1.04 (0.55 to 1.95) | - |
| **ER status** |  |  |
| Negative | Ref. | - |
| Positive | 0.97 (0.65 to 1.45) | - |
| **PR status** |  |  |
| Negative | Ref. | - |
| Positive | 0.93 (0.64 to 1.36) | - |
| **Her2 status** |  |  |
| Negative | Ref. | Ref. |
| Positive | 1.32 (0.87 to 1.99) | 1.12 (0.68 to 1.86) |
| **Breast histology** |  |  |
| Others | Ref. | Ref. |
| IDC | 0.61* (0.38 to 0.97) | 0.66 (0.37 to 1.19) |
| DCIS | 0.73 (0.29 to 1.82) | 0.36 (0.45 to 2.97) |
| ILC | 1.08 (0.55 to 2.12) | 1.79 (0.79 to 4.09) |
| **Chemotherapy** |  |  |
| No | Ref. | Ref. |
| Adjuvant | 0.57 (0.31 to 1.02) | 0.65 (0.30 to 1.38) |
| Neoadjuvant | 0.40* (0.16 to 0.97) | 0.41 (0.13 to 1.29) |
| **Radiotherapy (RT)** |  |  |
| No | Ref. | - |
| External RT | 0.69 (0.28 to 1.70) | - |
| IORT Boost | 1.51 (0.50 to 4.48) | - |
| IORT Radical | 0.79 (0.18 to 3.35) | - |
| **lymph vascular invasion (LVI)** |  |  |
| Negative | Ref. | - |
| Positive | 0.95 (0.63 to 1.43) | - |
| **Family history** |  |  |
| No | Ref. | Ref. |
| Second degree | 1.59* (1.01 to 2.51) | 1.66 (0.96 to 2.88) |
| First degree | 0.84 (0.44 to 1.58) | 0.96 (0.49 to 1.89) |
| **Lymph node metastasis** |  |  |
| 0 | Ref. | Ref. |
| 1~ | 0.65 (0.40 to 1.05) | 0.57 (0.27 to 1.20) |
| 4~ | 0.93 (0.56 to 1.52) | 0.36 (0.16 to 0.81) |
| >10 | 0.94 (0.49 to 1.78) | 0.30 (0.09 to 0.93) |
| **Stage** |  |  |
| I | Ref. | - |
| II | 0.98 (0.61 to 1.56) | - |
| III, IV | 0.88 (0.53 to 1.44) | - |
| **Grade** |  |  |
| I | Ref. | - |
| II | 0.89 (0.48 to 1.63) | - |
| III | 0.95 (0.50 to 1.8) | - |
| **Recurrence** |  |  |
| No | Ref. | Ref. |
| Yes | 2.17* (1.50 to 3.12) | 2.04* (1.25 to 3.33) |

\* $P < 0.05$

In multivariable analysis after adjusting for covariates, some variables had a significant association with CBC. The 60-70 years old patients (HR = 0.33; 95% CI: 0.11–0.92), 4-10 lymph node metastasis (HR = 0.36; 95% CI: 0.16–0.82), >10 lymph node metastasis (HR = 0.30; 95% CI: 0.09–0.94), recurrence (HR = 2.03; 95% CI: 1.24–3.13), were associated with developing CBC.

## Discussion

Although extensive research on breast cancer has led to successful treatment and survival of patients, the development of contralateral breast cancer after treatment of primary breast cancer has remained a major issue and needs further research.

In our study, the median time interval between PBC and CBC was 7.10 years which is to some extent consistent with elsewhere. In a study from Chicago, the median time interval between a PBC and CBC was reported as 6.2 years(16), and a different study from Sweden reported a median time interval of 6.7 years(17). The median time interval between PBC and CBC ranged from 3.4 to 5.6 years(18) (19). The incidence of CBC in our study was 3% after 5 years. This rate in other studies was 41.5% (16).

Nichols et al in a study on SEER data noted that early and late peak disease incidence age near 30 and 70 years, respectively(20). In a study by Lizarraga, I. M., et al. women younger than 35 years old at diagnosis were at higher risk for CBC(21). In another study by Liederbach, E., et al. patients ≥70 years old are at higher risk than patients younger than 50 years for CBC (16). The young cases (under 60 years old) are more related to CBC which maybe more related to amount of young people who are affected by BC in our country. In our study, in the age of less than 50 years old it was significantly higher than older.

In the present study family history of BC in the second-degree led to enhance the risk of CBC by 59%. The family history of BC among first-degree was not associated with an increased risk of developing CBC. Our finding is consistent with results from Yoon, T. I., et al and Vichapat, V., et al (22, 23). In other studies, investigators stated that women who have strong family history seem to be at higher risk of contralateral BC and they have cited family history as a potential risk factor for CBC(21, 24).

Some studies have shown that HER2^+^ has a significant association with overexpressing tumors(22, 25), but some else have not shown any association between contralateral BC and HER2^+^ (26, 27). In a study by Mruthyunjayappa, S., et al.(18) there was an association between bilateral BC and HER2-positive with a significantly lower rate. Although, no association was observed in the present study.

Studies that analyze metachronous BC risk factors most of the time adjust for the recurrence. In a study, investigators found that prior recurrence at a younger age has an increased risk of MBC(23). Our results show recurrence has a different effect on the development of CBC. The risk of CBC in patients with a recurrence of breast cancer is 2.03-fold.

In our study, radiotherapy hadn’t had a statistically significant effect on the incidence of CBC which is consistent with findings of some other studies (28, 29). Of course, this is related to kind of surgery as mastectomy or saving the breast. Hence, it’s not scientific statement around irradiation effectiveness. In Yadav, B. S., et al study(24), adjuvant chemotherapy hadn’t significant effect on the risk of second malignancy. In Alkner, S., et al. chemotherapy given after BC made a difference in the progression of disease and DFI. In this study, chemotherapy after BC was a negative prognostic factor(17). In the present study, adjuvant or neoadjuvant chemotherapy was not significantly protective or promotive for the CBC.

Although invasive lobular carcinoma is the second prevalent type of BC pathology(30), there are still conflicting results about the effect of invasive lobular carcinoma on CBC and it is controversial. According to Tong, J., et al study(31), patients who have invasive lobular carcinoma or a mixture of both compared to IDC alone BC have more likely to develop CBC but a study from the UK found that there was no significant difference between occurrence and time to occurrence of CBC in terms of pathology type(30). Glas et al revealed a higher risk of CBC in patients with lobular tumor morphology(32). In our study the ILC was 10.14% vs 5.78% in CBC and PBC groups, respectively and IDC was a protective factor for CBC.

In our study, most patients had lower lymph node metastasis. As the number of positive lymph nodes increased, the risk of CBC decreased which may be due to the worst outcome of them. This finding of the current study is consistent with those of Liederbach, E. et al. who found that CBC tend to be node-negative (16). The results of another study indicate that a lower rate of node-positive was significantly associated with metachronous bilateral BC(18).

The present study was performed for the first time in the area under work and has employed data from long-term follow-up of 20 years. This study has some limitations. First, the lack of access to some demographic variables such as reproductive and death criteria to analyze on those subgroups and adjust for those potential confounding risk factors. Outcome results of the CBC cases is not clearly in our study and should be into more consideration for researchers. The reason for the observed differences between our results and other studies could be due to differences in sample size and study settings.

## Conclusion

In the present study, CBC is prevalent in 2.9% of the BC patient’s survivors. This situation is more common in non-IDC cases (ILC) involved with criteria as younger age. These criteria are more related to induce CBC.

## Data Availability

All data produced in the present study are available upon reasonable request to the authors.

## Conflicts of interest

All authors declare that they have no conflict of interest about this study.

## Acknowledgments

The present research article has been extracted from a research project funded by the Breast Cancer Research Center at Shahid Beheshti University of Medical Sciences, Tehran, Iran.

## Notes

### Competing Interest Statement

The authors have declared no competing interest.

### Funding Statement

This study did not receive any funding

### Author Declarations

Ethics committee/1400.048 of Cancer Research Center, Shahid Beheshti Medical University gave ethical approval for this work.

### Summary of Updates

Determining the corresponding author

